# International Delphi Consensus on Risk Factors Justifying Antibiotic Prophylaxis in Transperineal Prostate Biopsy

**DOI:** 10.1101/2025.07.15.25331517

**Authors:** FP Stangl, F Wagenlehner, J Kranz, TE Bjerklund Johansen

## Abstract

**Background:** Transperineal prostate biopsy (TP-Bx) is increasingly favored over the transrectal approach due to its lower rate of infectious complications. Emerging evidence suggests that TP-Bx can often be safely performed without antibiotic prophylaxis in low-risk patients. However, there is no international consensus on the clinical risk factors that justify prophylactic antibiotic use, leading to wide practice variability and concerns regarding antimicrobial stewardship.

**Objective:** This Delphi consensus study aims to identify and define procedural, patient-related, and microbiological risk factors that warrant antibiotic prophylaxis in TP-Bx and to establish evidence-informed best practice recommendations.

**Methods:** A modified three-round Delphi process will be conducted in accordance with ACCORD and CREDES guidelines. A diverse panel of 50–60 international experts in urology, infectious diseases, microbiology, and antimicrobial stewardship will anonymously evaluate predefined and newly suggested risk factors using a 6-point Likert scale. Consensus will be defined as ≥70% of participants rating an item as highly important (score 5–6) and <15% rating it as unimportant (score 1–2). Items failing to reach consensus in Rounds 1 and 2 will be re-evaluated in subsequent rounds.

**Outcomes:** The primary outcome will be a consensus-based list of risk factors justifying antibiotic prophylaxis in TP-Bx. Secondary outcomes include recommendations for diagnostic testing, prophylactic regimens, and thresholds for initiating routine prophylaxis based on institutional infection rates.

**Dissemination:** Findings will be submitted for publication in peer-reviewed journals to support global efforts in harmonizing practice and promoting responsible antibiotic use in prostate cancer diagnostics.

## Background

Transperineal prostate biopsy (TP-Bx) is increasingly preferred over the traditional transrectal approach due to its lower rate of infectious complications. Recent high-quality evidence indicates that TP-Bx may be safely performed without antibiotic prophylaxis in low-risk patients^1-4^. However, there is currently no international consensus defining which risk factors that justifies periprocedureal prophylaxis. This lack of guidance has led to inconsistent practices worldwide, undermining both patient safety and antimicrobial stewardship. While antibiotic prophylaxis may reduce the risk of infectious complications, indiscriminate use can contribute to antimicrobial resistance and disruption of the microbiome—highlighting the critical balance between individual patient protection and broader antimicrobial stewardship principles^5-7^. To address this evidence-to-practice gap, we propose a Delphi consensus study to define a globally applicable set of clinical risk factors that justify antibiotic prophylaxis and provide a best practice recommendation for TP-Bx.

## Objectives

To identify and reach expert consensus on procedural-, patient- and microbiological risk factors that justify antibiotic prophylaxis and do define best practice in patients undergoing transperineal prostate biopsy.

## Study Design

We will employ a modified Delphi method consisting of three sequential and anonymous rounds. The study will adhere to the ACCORD (Accurate Consensus Reporting Document) and CREDES (Conducting and Reporting of Delphi Studies) guidelines to ensure transparency, methodological rigour, and reproducibility. The full protocol will be made publicly available before commencement of the study.

## Expert Panel

We aim to recruit approximately 50-60 experts in the fields of

- Urology
- Infectious Diseases and Antimicrobial Stewardship
- Microbiology

Participants will be selected based on

- Peer-reviewed publications
- Involvement in guideline panels
- Recognised clinical or academic leadership
- Personal experience with TP Bx (requirement for urologists only)

Experts will be drawn from across Africa, Asia, Australasia, Europe, Latin America, and North America to ensure global representation. In total 65 Experts will be invited to participate in the Delphi consensus. They will be contacted by email and two reminders will be sent as needed.

## Anonymity and Data Collection

All rounds will be conducted online using a secure survey platform managed by an external provider. Responses will be anonymised to prevent dominance bias and to allow candid input.

### Round 1

#### Participants will receive a structured survey comprising

A list of risk factors derived from an extensive literature review and from already established frameworks such as ORENUC, as well as recent Delphi consensus work on complicated urinary tract infections^8,9^ — including, for example, recent UTI, immunosuppression, diabetes, and prior sepsis after biopsy.

- Each item will be rated on a 6-point Likert scale, where
- 1–2 = Low or negligible importance
- 3–4 = Intermediate importance
- 5–6 = High importance

There will be open-text fields allowing participants to suggest additional risk factors not listed in the first round.

### Round 2

Participants will receive anonymised summary statistics from Round 1 (median, IQR, distribution). All original and newly suggested items will be re-rated, with the opportunity to revise scores based on group trends. Newly suggested items from round 1 will be presented.

### Round 3

Only items that did not reach a consensus in Round 2 will be presented. Participants will again rate each item on the 6-point scale, aiming to resolve disagreement.

### Consensus Definition

- Consensus in: ≥70% rate 5–6 and <15% rate 1–2
- Consensus out: ≥70% rate 1–2 and <15% rate 5–6
- No consensus: anything else

### Data Management and Analysis

All data will be processed anonymously. Summary statistics (medians, IQRs, consensus proportions) will be used for analysis. Answers will be stratified according to registered variables such as medical specialty, level of expertise, geographical representation etc. throughout the domains to provide a comprehensive reporting of all studied aspects related to best practice and antibiotic prophylaxis in transperineal biopsy. Free-text data will be coded thematically. Free-text data will be coded thematically.

## Ethics

This study involves no patients and poses no risk. As such, ethics approval is not required under applicable guidelines.

## Funding

The study is not funded but the platform for the Delphi Consensus (Within3) will be provided via an unrestricted grant by Advanz Pharma.

## Dissemination

The final consensus statement will be submitted for publication in high-impact journals.

### Timeline

- Month 1: Panel recruitment and item generation
- Months 2–3: Round 1
- Months 3–4: Round 2
- Months 4–5: Round 3
- Months 5–6: Analysis and manuscript preparation

Evidence background for suggested risk factors

| Domain | Risk Factor | Supporting Evidence / Source |
| --- | --- | --- |
| Procedural | Biopsy cores $\geq 20$ | More cores = higher tissue trauma and infection risk<sup>2</sup> |
|  | Repeat biopsy within 6 months | Tissue disruption and prior exposure can increase infection risk<sup>10</sup> |
|  | Simultaneous endoscopic/urologic procedures | Multiple manipulations increase contamination potential<sup>11</sup> |
|  | Prolonged procedure duration (>30 min) | Longer exposure correlates with higher SSI risk in surgical settings<sup>12</sup> |
|  | Absence of local antisepsis (e.g., iodine prep) | Standard in surgical infection prevention<sup>13</sup> |
|  | Inexperienced operator / early learning phase | More complications documented during training (supported in surgical simulation literature among different procedures)<sup>14</sup> |
|  | Hair Removal | Clipper causes less skin trauma<sup>15</sup> |
| Patient | Immunosuppression (e.g., steroids, chemotherapy) | Increased SSI risk in immunocompromised patients across surgical fields; extrapolated to TP biopsy<sup>16</sup> |
|  | Diabetes mellitus | Multiple studies show increased post-biopsy infection risk in diabetic patients<sup>17</sup> |
|  | Prostate Volume | Increased risk of post-biopsy retention possibly facilitating infection<sup>18</sup> |
|  | Indwelling urinary catheter | Risk of bacteriuria and UTI higher with catheters<sup>19</sup> |
|  | Recurrent urinary tract infections (≥2/year) | Historical UTI is a known predictor of recurrence and complications<sup>20</sup> |
|  | History of post-biopsy infection or urosepsis | Observational evidence supports recurrence risk<sup>21</sup> |
|  | Chronic prostatitis / CPPS | Increased inflammation and microbial presence may increase post-biopsy complications<sup>22,23</sup> |
|  | Recent hospitalization or institutionalization | Associated with MDRO colonization and higher infection rates<sup>24</sup> |
|  | Recent antibiotic use (last 3 months) | Promotes resistance and increases failure risk of standard prophylaxis<sup>25,26</sup> |
|  | Age >75 years | Older adults show increased infection and sepsis susceptibility<sup>27,28</sup> |
|  | ECOG ≥2 / Frailty | Functional impairment is associated with worse surgical outcomes<sup>29</sup> |
|  | Morbid obesity BMI >40 | <sup>30,31</sup> |
| **Microbiological** | Patient colonized with MDROs within 12 weeks before the procedure | Well-documented risk factor in multiple fields, including transrectal biopsy<sup>24,32</sup> |
|  | Local i.e. regional or if available hospital-wide high resistance rates (esp. FQ resistance) | Local ecology influences prophylaxis efficacy<sup>33</sup> |
|  | Prior isolation of ESBL-producing organisms (<6 Months) | Higher risk of failure of standard regimens<sup>34</sup> |
|  | Hospital-wide ongoing outbreak or clustering of urosepsis cases | Environmental risk<sup>35</sup> |

## Delphi Questionnaire

### Best Practice and Risk Factors for Antibiotic Prophylaxis in TP Biopsy

#### Procedural Factors

1. To what extent do you consider taking ≥20 biopsy cores a risk factor justifying antibiotic What is your medical speciality?

#### Please select answers (choose all that apply)

- Urologist
- Infectious disease specialist
- Microbiologist/ clinical microbiologist

2. How would you grade your own qualifications/knowledge related to the following: Please grade your level of expertise on a scale of 1-6 (1 means foundational/basic knowledge; 6 indicates expert knowledge)

- Prostate cancer treatment
- Prostate cancer diagnostics
- Prevalence and prevention of hospital acquired urogenital infections
- Antibiotic treatment of urogenital infections including sepsis
- Types of microorganisms and rates of resistance among pathogens causing urogenital infections
- Antimicrobial stewardship

3. What percentage of biopsies in your department are done with the following approach/guidance?

Please use the dropdown list to select an approximate percentage (to the nearest 5%) for each biopsy type. The total of all 3 percentages should be as close to 100% as possible. Any additional information can be added to the free-text box below. Once complete, please click the ‘Submit’ button.

- Transperineal ultrasound-guided biopsies
- Transrectal ultrasound-guided biopses
- Transperineal in-bore prostate biopsy

4. What percentage of targeted biopsies in your department are done with the following technique?

Please use the dropdown list to select an approximate percentage (to the nearest 5%) for each biopsy type. The total of all 3 percentages should be as close to 100% as possible. Any additional information can be added to the free-text box below. Once complete, please click the ‘Submit’ button.

- Image fusion technology (MRI and ultrasound)
- Cognitive guidance (Based on MRI image)

5. How many patients are typically scheduled per biopsy session (working day) in your centre?

- 0
- 1-3
- 4-6
- 7-9
- 10-11
- 12-14
- 15-16
- 17-19
- 20+

6. What is the average time allocated per TP biopsy procedure in your centre?

- 0-15 minutes
- 16-30 minutes
- 31-45 minutes
- 46-60 minutes
- 61-75 minutes
- 76-90 minutes
- >91 minutes

7. Is the biopsy performed as an outpatient, a day surgery, or an inpatient procedure?

- Outpatient
- Day surgery
- Inpatient

8. How many and what type of health personnel are typically involved in the operating theatre/outpatient room during a transperineal biopsy, in addition to the urologist who performs the procedure?

Please select the number next to each type of health personnel below

- A second urologist/ urology resident
- Radiologists
- Nurses
- Nurse’s aides
- Anesthesiologists
- Anesthesia nurse
- Other, please specify below

9. Who typically marks the biopsy targets on MRI images? Please select an answer:

- Radiologist
- Urologist
- Both together

10. Is hair removal routinely performed before transperineal prostate biopsy?

- Yes
- No

11. If hair removal is routinely performed before transperineal prostate biopsy, by whom is it removed?

- Patient
- Nurse
- N/A (if answered ‘No’ to Question 8)

12. If hair removal is routinely performed before transperineal prostate biopsy, where is it removed?

- Patient’s home
- Operating theatre
- N/A (if answered ‘No’ to Question 8)

13. If hair removal is routinely performed before transperineal prostate biopsy, how is it removed?

- Razor
- Clippers
- Hair removal cream
- N/A (if answered ‘No’ to Question 8)

14. If hair removal is routinely performed before transperineal prostate biopsy, when is it removed?

- Before the procedure
- As part of the procedure
- N/A (if answered ‘No’ to Question 8)

15. How is skin disinfection of the biopsy field performed?

- 2% chlorhexidine isopropanol solution
- Povidone iodine solution
- Benzoyl peroxide (BPO) gel
- Other

16. How is disinfectant applied?

- Sterile gauze
- Spray
- Other

17. Who typically administers the anaesthesia?

- Anaesthesist
- Urologist
- Nurse
- Other

18. What type of anaesthesia is typically used in your centre?

#### Please select answers (choose all that apply)

- Local
- Sedation
- General
- Spinal

19. Do you routinely administer antibiotic prophylaxis in transperineal biopsy?

#### Please select an answer

- Yes
- No
- In high risk patients only (to be defined in this consensus)

20. What route of antibiotic prophylaxis is typically used in your centre?

- None
- Oral
- Intravenous
- Intramuscular

21. What type of antibiotics do you use?

#### Please select answers (choose all that apply)

- 1^st^ generation cephalosporins (e.g., Cefazoline)
- 2^nd^ generation cephalosporins (e.g., Cefuroxime)
- 3^rd^ generation cephalosporins (e.g., Ceftriaxone)
- Fosfomycin
- Aminoglykosides (e.g., Gentamicin)
- Aminopenicillins (e.g., Amoxicillin)

22. What is your main rationale for the choice of antibiotic prophylaxis in transperineal prostate biopsy?

#### (Please select all that apply.)

- To cover skin flora (e.g., Staphylococcus spp., gram-positive cocci)
- To provide broad-spectrum coverage, including gram-negative organisms
- To target uropathogens commonly found postbioptic prostatitis
- To align with recommendations for other clean-contaminated urologic procedures
- Based on personal experience with post-biopsy infections
- No specific microbiological rationale — prophylaxis according to routine in-hospital guideline
- Other (please specify): ___________

23. Do you routinely obtain a urine culture before transperineal biosy to assess asymtpomatic bacteriuria (ABU)?

#### Please select an answer

- Yes
- No

24. In case of ABU, do you administer additional antibiotics? Please select an answer:

- Yes
- No

25. If you answered ‘Yes’ to ‘In case of ABU, do you administer additional antibiotics?’

- A short treatment course before the procedure
- Periprocedural prophylaxis only
- Both preprocedural treatment and periprocedural prophylaxis
- N/A (answered ‘No’ to the previous question)

26. Are you monitoring the detection rate of prostate cancer with transperineal prostate biopsy?

- Yes
- No

27. How many transperineal biopsy procedures or transperineal needle-based prostate treatment procedures have you done/supervised yourself (Approximately)?

- Please provide an approximate number

28. Have you ever seen infections after transperineal biopsy?

- Yes
- No
- If yes,

#### Please write the type of infection (s)

#### Please write the type of pathogen(s)

If your observations have been published, please provide the reference

29. What type of access do you use for TPBx?

#### Please select an answer

- Template-guided access
- Single-entry port
- Other, please specify below

30. Please describe your biopsy sampling methods

#### Please select answers (choose all that apply)

- Systematic biopsy without targeted biopsy
- Systematic biopsy with targeted biopsy
- Targeted biopsy only
- Targeted biopsy and perilesional sampling

31. How many cores are usually taken in one biopsy procedure for a systematic biopsy only? Please provide the number (or range) for:

- Critical volume for when extra cores are taken (e.g., 50cc):
- Number of extra cores per volume unit (e.g., 1 extra core per 10cc):

32. How many cores are usually taken in one biopsy procedure for a systematic biopsy with targeted biopsy?

#### Please provide the number (or range) for

- Number of cores per target lesion (>PIRADS 3):

33. How many cores are usually taken in one biopsy procedure for a targeted biopsy only?

- Please provide the number (or range) for:
- Number of cores per target lesion (>PIRADS 3):
- Number of cores per exact (> PIRADS 3) anatomic location (e.g., apical bilaterally, posteromedial bilaterally):

34. How many cores are usually taken in one biopsy procedure for a targeted biopsy and perilesional sampling?

#### Please provide the number (or range) for

- Number of perilesional cores per target (PIRADS 3 or above):

35. In which contamination category will you place transperineal prostate biopsy?

- Please select an answer:
- Clean
- Clean contaminated (Urinary tract)
- Clean contaminated (Bowel)
- Contaminated

37. If a urology department monitored the overall rate of infective complications without prophylaxis (including all risk factors), at which level of infective complications would you recommend starting routine antibiotic prophylaxis?

#### (Please tick your lowest acceptance rate only) Please select an answer

- 1%
- 2%
- 3%
- 4%
- 5%
- 10%
- 15%
- 20%

### Patient-Related Factors

38. To what extent do you consider that the risk factors below justify antibiotic prophylaxis in TP biopsy?

- Please use the dropdown list to rate each risk factor via the following 6-point Likert scale.

Any additional information can be added to the free-text box below. Once complete, please click the ‘Submit’ button before progressing to the next question.

#### Likert Scale

- 1–2 = Low or negligible importance
- 3–4 = Intermediate importance
- 5–6 = High importance
- If you feel you do not have the necessary expertise to answer the questions is this domain, please proceed to Question 3
- Please rate the following:
- Taking ≥20 biopsy cores
- Performing a repeat biopsy within 6 months
- Performing a simultaneous endoscopic/urologic procedure
- Procedure duration >30 minutes
- Violation of sterility in the surgical field (e.g., poor margins/coverage, iodine prep.)
- Inexperienced operator
- Please feel free to elaborate on your response

39. If two or more of the below risk factors are present, would you consider that the risk factors below justify antibiotic prophylaxis in TP biopsy?

- Taking ≥20 biopsy cores
- Performing a repeat biopsy within 6 months
- Performing a simultaneous endoscopic/urologic procedure
- Procedure duration >30 minutes
- Violation of sterility in the surgical field (e.g., poor margins/coverage, iodine prep.)
- Inexperienced operator
- Please select an answer:
- Yes
- No
- Depends, please elaborate below

40. If you feel you do not have the necessary expertise to answer the questions in this domain, please answer below

- Please select an answer:
- I do not have the necessary expertise to answer the questions in this domain
- I am confident in this domain and have answered the questions

41. To what extent do you consider that the risk factors below justify antibiotic prophylaxis in TP biopsy?

- Please use the dropdown list to rate each risk factor via the following 6-point Likert scale. Any additional information can be added to the free-text box below. Once complete, please click the ‘Submit’ button before progressing to the next question.

#### Likert Scale

- 1–2 = Low or negligible importance
- 3–4 = Intermediate importance
- 5–6 = High importance

If you feel you do not have the necessary expertise to answer the questions is this domain, please proceed to Question 5

- View the attached resources below (ECOG and FRAILTY scores)
- Please rate the following:
- Diabetes mellitus (poorly regulated or in need of medical treatment)
- Immunosuppression (e.g., steroids, chemotherapy) prophylaxis in TP biopsy?
- The presence of an indwelling urinary catheter
- Recurrent UTIs (≥2/ 6 months or > 3/ 1 year)
- A history of post-biopsy infection or urosepsis
- A history of chronic prostatitis / CPPS
- Hospitalization or other institutionalization within the recent 3 months
- Antibiotic usage within the recent 3 months
- Age >75 years
- ECOG ≥2
- Morbid obesity (BMI ≥40)
- Critical frailty score ≥4

42. If two or more of the below risk factors are present, would you consider that this situation justifies antibiotic prophylaxis in TP biopsy?

- Diabetes mellitus (poorly regulated or in need of medical treatment)
- Immunosuppression (e.g., steroids, chemotherapy) prophylaxis in TP biopsy?
- The presence of an indwelling urinary catheter
- Recurrent UTIs (≥2/ 6 months or > 3/ 1 year)
- A history of post-biopsy infection or urosepsis
- A history of chronic prostatitis / CPPS
- Hospitalization or other institutionalization within the recent 3 months
- Antibiotic usage within the recent 3 months
- Age >75 years
- ECOG ≥2
- Morbid obesity (BMI ≥40)
- Critical frailty score ≥4

### Microbiological Factors

43. Would any of the risk factors below justify pre-biopsy urine culture, rectal swab culture, or perineal skin swab culture?

- Diabetes mellitus (poorly regulated or in need of medical treatment)
- Immunosuppression (e.g., steroids, chemotherapy) prophylaxis in TP biopsy?
- The presence of an indwelling urinary catheter
- Recurrent UTIs (≥2/ 6 months or > 3/ 1 year)
- A history of post-biopsy infection or urosepsis
- A history of chronic prostatitis / CPPS
- Hospitalization or other institutionalization within the recent 3 months
- Antibiotic usage within the recent 3 months
- Age >75 years
- ECOG ≥2
- Morbid obesity (BMI ≥40)
- Critical frailty score ≥4

44. Which pathogen species will you consider when deciding on an antibiotic prophylaxis regimen in transperineal prostate biopsy?

- Please select an answer:
- Gram negative
- Gram positive
- Others, please specify below

45. If you feel you do not have the necessary expertise to answer the questions in this domain, please answer below

- Please select an answer:
- I do not have the necessary expertise to answer the questions in this domain
- I am confident in this domain and have answered the questions

46. To what extent do you consider that the risk factors below justify antibiotic prophylaxis in TP biopsy?

Please use the dropdown list to rate each risk factor via the following 6-point Likert scale. Any additional information can be added to the free-text box below. Once complete, please click the ‘Submit’ button before progressing to the next question.

#### Likert Scale

1–2 = Low or negligible importance

3–4 = Intermediate importance

5–6 = High importance

If you feel you do not have the necessary expertise to answer the questions is this domain, please proceed to Question 4

- Please rate the following:
- Patient is colonized with MDR microorganisms (Any sample)
- --Select Option --
- Isolation of ESBL-producing organisms in patient within recent 6 months (Any sample)
- A hospital-wide outbreak or clustering of urosepsis cases

47. If two or more of the below risk factors are present, would you consider that the risk factors below justify antibiotic prophylaxis in TP biopsy?

- Patient is colonized with MDR microorganisms (Any sample)
- Isolation of ESBL-producing organisms in patient within recent 6 months (Any sample)
- A hospital-wide outbreak or clustering of urosepsis cases Please select an answer:
- Yes
- No
- Depends

48. Would any of the below risk factors justify pre-biopsy urine culture or rectal swab culture?

- Patient is colonized with MDR microorganisms (Any sample)
- Isolation of ESBL-producing organisms in patient within recent 6 months (Any sample)
- A hospital-wide outbreak or clustering of urosepsis cases Please rate the following:
- Urine culture
- Rectal culture

49. If you feel you do not have the necessary expertise to answer the questions in this domain, please answer below

- I do not have the necessary expertise to answer the questions in this domain
- I am confident in this domain and have answered the questions

#### Final Remarks

50. Thank you for answering the questions. Please use this space to ask any questions or provide any additional information.

## Data Availability

All data produced in the present study are available upon reasonable request to the authors

## Notes

### Competing Interest Statement

The authors have declared no competing interest.

